# The German Corona Consensus Dataset (GECCO): A standardized dataset for COVID-19 research

**DOI:** 10.1101/2020.07.27.20162636

**Authors:** Julian Sass, Alexander Bartschke, Moritz Lehne, Andrea Essenwanger, Eugenia Rinaldi, Stefanie Rudolph, Kai U. Heitmann, Jörg J. Vehreschild, Christof von Kalle, Sylvia Thun

## Abstract

**Background:** The current COVID-19 pandemic has led to a surge of research activity. While this research provides important insights, the multitude of studies results in an increasing segmentation of information. To ensure comparability across projects and institutions, standard datasets are needed. Here, we introduce the “German Corona Consensus Dataset” (GECCO), a uniform dataset that uses international terminologies and health IT standards to improve interoperability of COVID-19 data.

**Methods:** Based on previous work (e.g., the ISARIC-WHO COVID-19 case report form) and in coordination with experts from university hospitals, professional associations and research initiatives, data elements relevant for COVID-19 research were collected, prioritized and consolidated into a compact core dataset. The dataset was mapped to international terminologies, and the Fast Healthcare Interoperability Resources (FHIR) standard was used to define interoperable, machine-readable data formats.

**Results:** A core dataset consisting of 81 data elements with 281 response options was defined, including information about, for example, demography, anamnesis, symptoms, therapy, medications or laboratory values of COVID-19 patients. Data elements and response options were mapped to SNOMED CT, LOINC, UCUM, ICD-10-GM and ATC, and FHIR profiles for interoperable data exchange were defined.

**Conclusion:** GECCO provides a compact, interoperable dataset that can help to make COVID-19 research data more comparable across studies and institutions. The dataset will be further refined in the future by adding domain-specific extension modules for more specialized use cases.

## INTRODUCTION

In December 2019, first reports of a cluster of 41 patients infected by a novel coronavirus emerged from Wuhan, China.^1^ Within a few months, the new virus, subsequently named “severe acute respiratory syndrome coronavirus 2” (SARS-CoV-2), has spread around the world causing the global COVID-19 pandemic. Currently (as of July 1, 2020), SARS-CoV- 2 has infected more than 10 million and killed more than half a million patients worldwide.^2^

The pandemic has spurred intensive scientific research, including numerous regional, national and international epidemiological surveys and studies.^3–7^ While this research provides important new insights, the multitude of studies threatens to generate a dangerous segmentation of information. This could delay or even prevent urgently needed scientific knowledge about SARS-CoV-2 and COVID-19. To avoid this segmentation of information and make COVID- 19 data more comparable and exchangeable across studies and institutions, interoperable datasets are needed.

Various initiatives have started to define uniform datasets and Common Data Elements (CDEs) for the collection of information about COVID-19. For example, questionnaires and case report forms (CRFs) have been developed to collect data about COVID-19 patients in a standardized way.^5,8,9^ While the CDEs defined in these projects are an important step, they are not enough to ensure interoperability. To make data syntactically and semantically interoperable, data elements also have to be embedded in standard data structures that can be exchanged across IT systems, and they have to use common terminologies that unambiguously define the meaning of clinical concepts.

To improve interoperability of COVID-19 data, we developed the German Corona Consensus Dataset (GECCO), which uses international health IT standards and terminologies for interoperable data exchange. GECCO defines a compact set of data elements to be collected in COVID-19 studies and was developed within the National Research Network of University Medicine on COVID-19 (“Nationales Forschungsnetzwerk [NFN] der Universitätsmedizin zu COVID-19”) funded by the German Federal Ministry of Education and Research (BMBF). The following paper provides an overview of the GECCO dataset and its development.

## METHODS

### Selection of data elements

An initial dataset was compiled as a working basis by merging data elements and response options of the following projects: the ISARIC-WHO CRF^8^; the Pa-COVID-19 study^10^, which investigates the pathophysiology of COVID-19 in a prospective patient cohort; the LEOSS case registry^3^, a clinical patient registry for patients infected with SARS-CoV-2 initiated by the ESCMID Emerging Infections Task Force (EITaF), the German Center for Infection Research (DZIF) and the German Society for Infectiology (DGI). This draft dataset was saved in a spreadsheet and sent to members of an expert board for comment and proposal of additional data elements. The expert board was composed of health professionals from German university hospitals, professional associations and other relevant organizations. New data elements proposed by the expert board were added to the dataset for subsequent prioritization. For the prioritization, the experts were asked to assign a priority value to each data element of the dataset. Priorities were indicated on a 5-level scale that was loosely based on the NIH model for CDEs^11^ (Table 1).

**Table 1:** Prioritization of data elements.

| Scale value | Priority | NIH Classification | Definition |
| --- | --- | --- | --- |
| 5 | highly relevant | General Core /<br>Disease Core* | Data element with essential general or specific information relevant to COVID-19 |
| 4 | very relevant | Supplemental –<br>Highly<br>Recommended | Data element that is essential under certain conditions or for certain study types and is therefore strongly recommended |
| 3 | relevant | Supplemental | Data element that is often collected in clinical studies, but whose relevance depends on the study design or type of research |
| 2 | less relevant | Exploratory | Data element that requires further validation, but which can fill current gaps in the data elements and/or replace an existing data element |
| 1 | not relevant | - | Data elements that are not considered relevant to the dataset |

*\* Since this is a disease-specific (i.e. COVID-19) dataset, both the general and disease-specific core categories of the NIH were assigned to the highest priority level.*

From the data elements with the highest prioritizations, a preliminary core dataset with roughly 100 data elements was compiled (this size was chosen to include as many relevant data elements as possible, while keeping the dataset manageable and practical). This core dataset was then reviewed by an editorial team of seven experts from different disciplines. In consensual decisions, data elements not considered necessary for the core dataset were discarded; conversely, data elements that were considered highly important but had not yet been included in the core dataset were added. The final data elements of the core dataset were grouped into meaningful categories (e.g., demographics, symptoms or medication). Figure 1 shows the workflow of consensus building and dataset definition.

**Figure 1:**
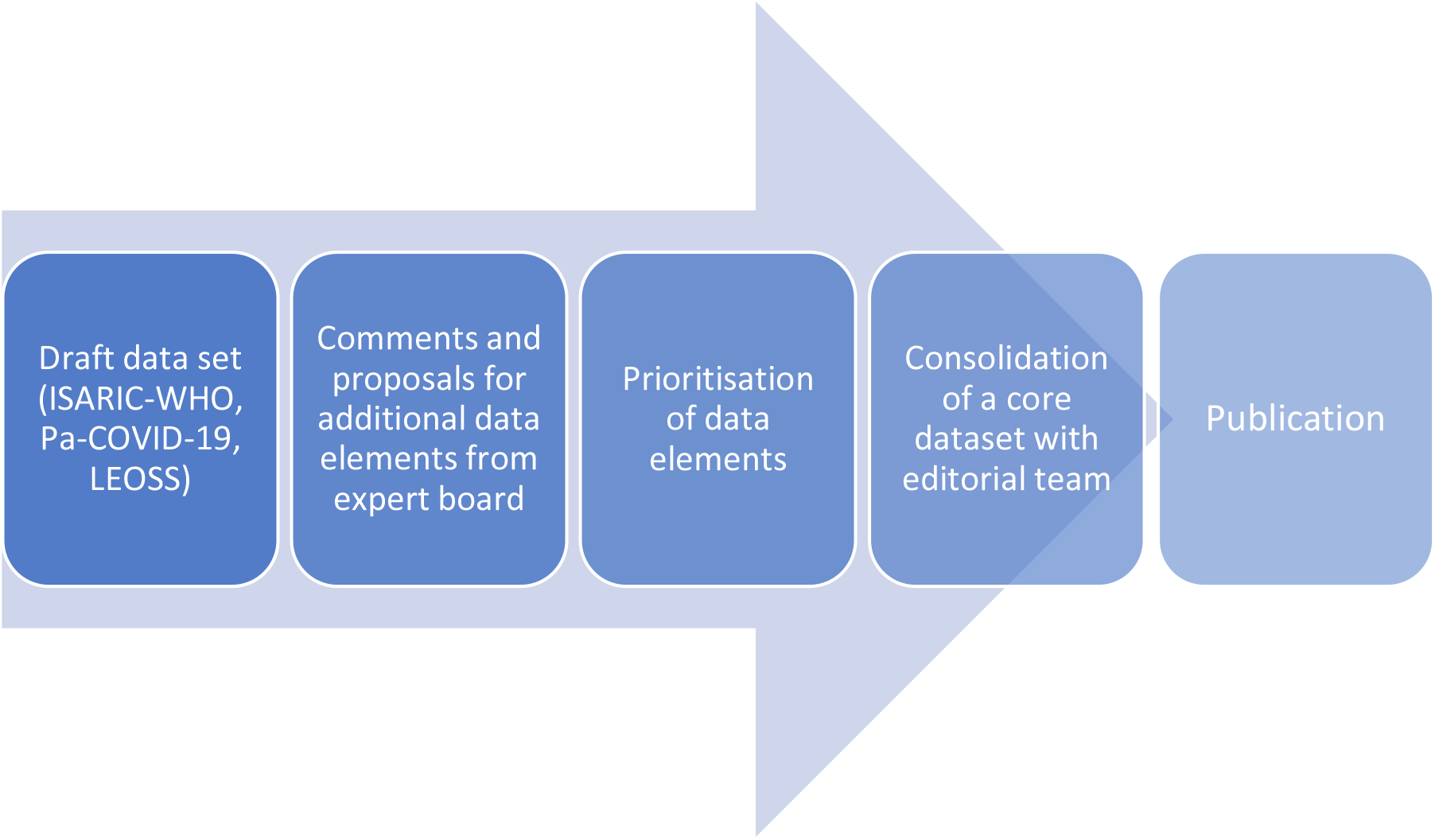
Workflow of consensus building and definition of data elements for the GECCO core dataset.

### Standardization

To ensure syntactic and semantic interoperability, elements and response options of the core dataset were mapped to international standards and terminologies. The following terminologies and code systems were used: the International Statistical Classification of Diseases and Related Health Problems, 10^th^ revision, German modification (ICD-10-GM)^12^ for diagnoses; Logical Observation Identifiers Names and Codes (LOINC)^13^ for laboratory values and other measurements; the Unified Code for Units of Measure (UCUM)^14^ for measurement units; the Anatomical Therapeutic Chemical Classification System (ATC)^15^ for active ingredients of drugs and medications; SNOMED CT^16^ for diagnoses and other medical concepts. The annotation of data elements with international terminologies was done using ART-DECOR^17^, an open source collaboration platform for experts from medical, terminological and technical domains aiming on creation and maintenance of datasets with data element descriptions, use case scenarios, value sets and Health Level 7 (HL7) templates and profiles.

To define interoperable formats for data exchange, the HL7 standard “Fast Healthcare Interoperability Resources” (FHIR)^18^ was used. FHIR builds on a set of “resources”, which provide generic data structures for common healthcare concepts, such as Patient, Practitioner, Observation, Medication or Condition. From these resources more specific data structure definitions, so-called “profiles”, can be defined, which allow for interoperable data exchange across health IT systems. To ensure interoperability, care was taken to build on previous work where possible, in particular the FHIR profiles of the German Medical Informatics Initiative^19^, the International Patient Summary (IPS)^20^, the Logica COVID-19 profiles^21^ and the FHIR base profiles of HL7 Germany.^22^ FHIR profiles were defined using Forge^23^ and published on the Simplifier platform.^24^

## RESULTS

Combining the initial draft dataset and the additional proposals from the expert board, 702 potentially relevant data elements were collected. From these data elements and based on the prioritization of the expert board, the editorial team compiled a core dataset consisting of 81 elements with 281 response options. These data elements were grouped into the following categories: anamnesis / risk factors (n = 16); imaging (n = 2); demographics (n = 7); epidemiological factors (n = 1); complications (n = 1); onset of illness / admission (n = 1); laboratory values (n = 25); medication (n = 4); outcome at discharge (n = 3); study enrollment / inclusion criteria (n = 2); symptoms (n = 2); therapy (n = 6); vital signs (n = 11) (Figure 2).

**Figure 2:**
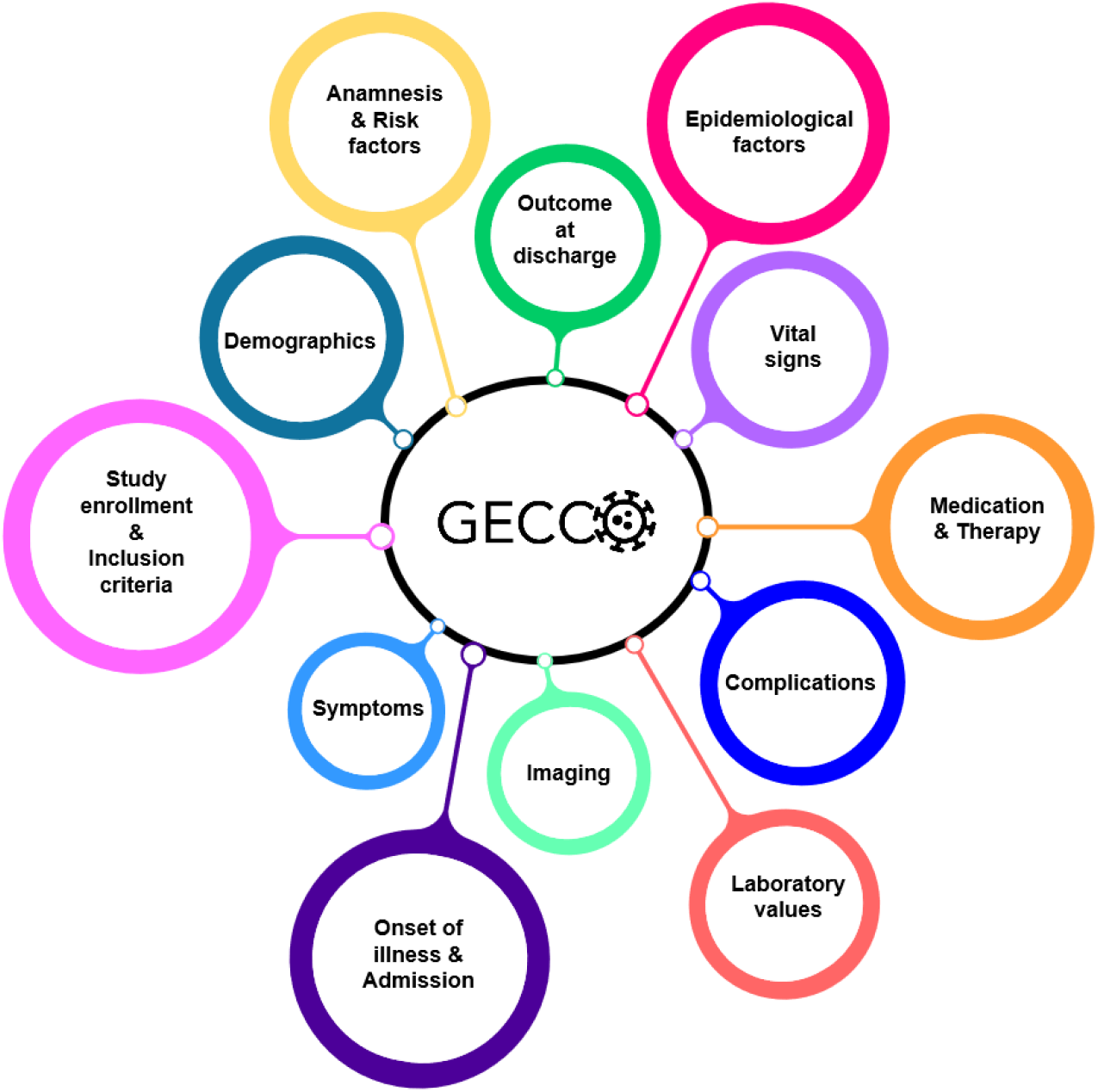
GECCO dataset categories into which data elements were grouped.

For all data elements and their corresponding response options, value sets were created using codes from SNOMED CT, LOINC, UCUM, ICD-10-GM and ATC. Data elements, response options and associated value sets can be accessed at https://art-decor.org/art-decor/decor-datasets--covid19f-.

Subsequently, FHIR profiles were created for the data elements. The following FHIR resources were used to model the data elements: Patient, Consent, Observation, Condition, Procedure, Encounter, Medication and MedicationStatement. The FHIR profiles can be accessed at https://simplifier.net/ForschungsnetzCovid-19.

During the consolidation process, it became clear that some data elements are important for certain disciplines but irrelevant for others. These elements were not included in the core dataset as they would have inflated the size of the dataset. The editorial team decided to include these data elements in domain-specific extension modules, which will be specified in more detail at later stages of the project.

## DISCUSSION

In this report, we presented the GECCO dataset, a core collection of data elements for acquiring and exchanging information about COVID-19 patients. By using standardized data structures (HL7 FHIR profiles) and international terminologies, the GECCO dataset is an important step towards interoperability of COVID-19 research data. It can facilitate harmonized data collection and analysis across institutions and IT systems, for example in clinical studies, registries or digital health applications.

A key factor to the successful application of standard datasets like GECCO is a close collaboration with the scientific community. To ensure a high acceptance of the dataset, the development of GECCO therefore included clinicians from a wide variety of medical disciplines and professional associations as well as experts in digital health, standardization and clinical terminologies. GECCO also collaborates closely with standards developing organizations such as HL7 and Integrating the Healthcare Enterprise (IHE) as well as other initiatives aiming to improve health data interoperability, such as the Medical Informatics Initiative^25^, NFDI4Health^26^ and the Corona Component Standards (cocos)^27^. Building on this strong consensus, GECCO-based data collection now has become a requirement for projects funded by the National Research Network of University Medicine on COVID-19 (NFN).

Although the GECCO dataset was designed to be as compact and manageable as possible, acquiring and recording the information for all data elements still requires time (for example, when entering the information in an electronic case report form). Moreover, manual documentation is prone to transcription errors. Conversely, manually abstracted and structured information from unstructured health records may provide relevant insights for care-providers and improve their understanding of risk and outcome. For some of the data items, it is therefore desirable to automatically exchange data between a GECCO-based study database and existing IT systems, such as hospital information systems or clinical trial software. This requires standard interfaces between these systems. The FHIR profiles of the GECCO dataset provide an interoperable, machine-readable data structure that can facilitate this data exchange across IT systems.

Scientific knowledge about COVID-19 and SARS-CoV-2 is changing fast, which may necessitate modifications to the GECCO dataset in the future. To incorporate new knowledge into the dataset, the NFN will put a governance framework in place that will coordinate revisions and extensions to the dataset. Domain-specific extension modules are already in preparation. Extension modules currently planned are: laboratory, diagnostics, immunology, gynecology and pregnancy, epidemiology, pediatrics, intensive care, oncology, radiology, virology, psychiatry and neurology (these extension modules are also accessible on the ART-DECOR platform).

## CONCLUSION

The GECCO dataset provides researchers and healthcare professionals with a compact, interoperable dataset for collecting, exchanging and analyzing COVID-19 data across institutions and software systems. Developed by a multidisciplinary group of experts, GECCO builds heavily on international terminologies and IT standards. GECCO can thus help to improve the harmonization and coordination of research efforts to successfully fight the COVID-19 pandemic. Future inclusion of domain-specific extension modules will further expand the use of the GECCO dataset.

## Data Availability

Data elements, response options, value sets and FHIR profiles of the GECCO dataset are publicly available on ART-DECOR and Simplifier (see links).

https://art-decor.org/art-decor/decor-datasets--covid19f-

https://simplifier.net/ForschungsnetzCovid-19

## ACKNOWLEDGMENTS

We thank the members of the expert board and editorial team for their help with the development of the GECCO dataset.

## DATA AVAILABILITY

Data elements, response options and value sets of the GECCO dataset can be accessed at https://art-decor.org/art-decor/decor-datasets--covid19f-. FHIR profiles are available at https://simplifier.net/ForschungsnetzCovid-19.

